# Development of tool for assessment of performance motivation and job satisfaction among the Community Health Workers of Central India

**DOI:** 10.1101/2022.01.10.22268956

**Authors:** G Revadi, Ankur Joshi, Abhijit P Pakhare

## Abstract

**Introduction:** Among the various factors influencing the performance of Community health workers, motivation and job satisfaction serves as a potential drive to perform better. Hence, this study aimed at constructing a motivation and job satisfaction tool in a systematic method that would serve as a potential tool for further research considering the heterogeneity in the study population.

**Objective:** To develop a tool to measure how well the CHWs are motivated and satisfied pertaining to individual, community and health system determinants.

**Methods:** This cross-sectional study from rural block of Madhya Pradesh in Central India included relatively high performing and low performing CHWs based on their annual performance-based incentives for the year (April 2017-March 2018). The CHWs were administered a self-reported questionnaire that contained a 5-point Likert scale with individual, health system and community determinants of motivation and job satisfaction.

**Results:** The performance motivation scale with 18 items and job satisfaction scale with 15 items were administered to the 92 CHWs. Their item content validity index was 0.66 and 0.83 respectively. The finalized tool consisted of 11 items in motivation scale and all the 15 items in job satisfaction scale following the Explanatory factor analysis. All the individual constructs in both the scales showed good internal consistency with Cronbachs alpha ranging from 0.62-0.88. The overall median (IQR) score of both RLP and RHP CHWs were 4(4–5) in both the questionnaires.

**Conclusion:** The CHWs in our study were intrinsically motivated and were satisfied with their performance as voluntary village health workers. Further research would be planned to validate the constructs using confirmatory factor analysis.

## Introduction

Community health workers (CHWs) are defined as those who play a defined role in the community and health system thereby acting as a bridge in providing primary health care services to the community (1). One of the key components of the National Health Mission in India is to provide every village with a population of 1000 in the country with a trained female community health activist known as ASHA i.e., Accredited Social Health Activists (2). A unique approach of what health system faces while delivering primary health services is to motivate the CHWs and to prevent attrition (3). Motivation refers to an individual’s degree of willingness to exert and maintain effort on assigned tasks. Satisfaction refers to which the CHWs derive personal satisfaction from serving the community, providing good quality services (4).

Tripathy et al. (5) showed that motivating factors of CHWs performance were financial incentives, passion for work, community recognition, trust and feeling of pride and training (5). Gopalan et al. (6) focused on the pattern of motivation where CHWs were better motivated over individual and community level factors when compared with the health system factors. Other enabling factors for motivation were better utilization of time, lack of other job opportunities, sense of community service, sense of holding government jobs and support from the peers for smooth functioning (7–9). Similarly, the demotivators include job insecurity, health problems in CHWs and heavy workload (5). The disabling factors were more towards the health system factors limiting their performance like organization of meetings and trainings at distant location of their village, resource constraint and lack of transport facility for referral (6).

Motivation and job satisfaction were considered as a feasible output measure of CHWs performance in the logic model (4) as these factors were a result of complex interaction of incentives, supervision, training, community involvement and co-ordination with the peer groups (10). Heterogeneity or variation of the above factors persists between Global and Indian studies. Madhya Pradesh being a high focus state with literature scarcity, it is crucial to understand and explore how well the CHWs are motivated and satisfied. Hence based on our literature search, motivation and job satisfaction questionnaire were shortlisted and we aimed to construct a questionnaire that serves as a potential tool for further research considering the heterogeneity.

## Methodology

This cross-sectional study was conducted with CHWs as study participants in a rural setting (Obedullaganj block) of Madhya Pradesh that included three Primary health care centres (PHCs) and three Community health care centres (CHCs). Data on performance-based incentives was retrieved from the Block Programme Management unit for the year (April 2017 – March 2018) which formed the basis for quantitative sampling plan. Out of 189 CHWs in the block, all those who received above 75^th^ percentile (arbitrarily termed as Relatively high performing, RHP) and below 25^th^ percentile (termed as Relatively low performing) of annual performance-based incentives in the last financial year were purposively included and those who were performing in the urban area of Obedullaganj block were excluded. This stratification was attempted to understand the level of motivation and job satisfaction of RHP and RLP CHWs.

## Data collection Methods

The finalized questionnaire in the XLS form was entered in ONA software and integrated to the mobile based ODK app. This Self-administered Questionnaire included demographic details and audio recording of how to fill Likert scale of motivation and job satisfaction questionnaire. Among the 189 CHWs, the 93 CHWs who fulfilled the criteria i.e, 46 RLP and 46 RHP CHWs agreed to participate and responded to the questionnaire. The selected CHWs were approached during their monthly meetings at their respective PHCs and village health and nutrition days (once or twice a month) through their facilitators and were briefed about the study questionnaire and their consent was obtained.

## Ethical clearance

Institutional Human Ethics Committee (IHEC) of AIIMS Bhopal, India, approved this study IHEC-LOP/2018/MD0027 Dated 9/10/2018. Permission and facilitation for data collection at field sites were provided by Block Medical Officer, Obedullaganj block, Raisen District, Madhya Pradesh.

## Tool development

The development of the motivation and the job satisfaction questionnaire for administering the CHWs involved a five-step process (11) where primarily those variables which influence the motivation (5,6,12) and job satisfaction (13) were reviewed from the literature and enlisted. Secondly the internal validity of the questionnaire (14) followed by pilot testing of the questionnaire, exploratory factor analysis and finally reliability analysis using internal consistency was done.

## Statistical analysis

Data in the form of excel sheet was imported from the Ona software and following data cleaning data analysis was done using IBM SPSS version 24 for data analysis. Nominal or categorical variables were summarized as frequency and percentage. Continuous variables were summarized as median and interquartile range as they were non-normally distributed. For each determinant, association of numerical variable with binary dependent outcome (RHP/RLP) was done using Mann-Whitney test. Performance Motivation Assessment Scale and job satisfaction scale was validated initially by Content Validity Index and then by Exploratory Factor Analysis (Principal Component Analysis). P value < 0.05 was considered significant.

## Operational definitions

### Rural area

Those places which do not satisfy the below criteria comes under the definition of rural (15).

- all places with a Municipality, Corporation or Cantonment or Notified Town Area
- a minimum population of 5000
- at least 75% of the male working population engaged in non-agricultural work.
- a density of population of at least 400 sq. Km.

#### Village

The smallest area of habitation in the rural area. It follows the limits of a revenue village that is recognized by the normal district administration. The revenue village need not necessarily be a single agglomeration of the habitations and accounts to 1 unit (15).

#### Block

Community development block is a rural area administratively earmarked for planning and development. A community development block covers several gram panchayats, the local administrative units at the village level (16).

#### PHCs

is a cornerstone for rural health services. A typical Primary Health Centre covers a population of 20,000 in hilly, tribal, or difficult areas and 30,000 populations in plain areas with 6 indoor/observation beds (17).

#### CHCs

The CHCs were designed to provide referral health care for cases from the Primary Health Centres level and for cases in need of specialist care approaching the centres directly. 4 PHCs are included under each CHC thus catering to approximately 80,000 populations in tribal/hilly/desert areas and 1,20,000 population for plain areas (18).

### Incentives

National Health Mission in India has developed a performance Based Incentives system wherein the ASHAs are paid according to the health activities they perform (19) e.g., Rs. 300 for registration of pregnancy.

### CHW facilitators

They are the main vehicle of monitoring, supportive supervision and on-site assistance for the CHWs. One ASHA facilitator is expected to approximately support 20 ASHAs (20).

## Results

The socio demographic details of the CHWs stratified according to their performance has been described in Table 1 among which the median (IQR) age of the CHWs were 30 (27-35) and the median (IQR) years of experience was 7(5-11).

**Table 1:** Distribution of Socio-demographic characteristics of CHWs (N =92)

| Sr<br>N0 | Variables | RLP n (%)<br>N=46 | RHP n (%)<br>N=46 | Total<br>N=92 |
| --- | --- | --- | --- | --- |
| 1 | Age |  |  |  |
|  | Less than or equal to 25 | 13(28.3) | 0 | 13 |
|  | More than 25 | 33(71.7) | 46(100) | 79 |
| 2 | Education |  |  |  |
|  | Up to Primary education | 25(54.3) | 14(30.4) | 39 |
|  | Others | 21(45.7) | 32(69.6) | 53 |
| 3 | Marital status |  |  |  |
|  | Married | 43(93.5) | 42(91.3) | 85 |
|  | Others | 3(6.5) | 4(8.7) | 7 |
| 4 | Religion |  |  |  |
|  | Hindu | 46(100) | 41(89.1) | 87 |
|  | Other religion | 0 | 5(10.9) | 5 |
| 5 | Caste |  |  |  |
|  | General | 13(28.3) | 6(13) | 19 |
|  | OBC | 17(37) | 17(37) | 34 |
|  | Scheduled Caste | 10(21.7) | 6(13) | 16 |
|  | Scheduled Tribe | 6(13) | 17(37) | 23 |
| 6 | Socio-economic status |  |  |  |
|  | Above poverty line | 15(32.6) | 21(45.7) | 36 |
|  | Below poverty line | 31(67.4) | 25(54.3) | 56 |
| 7 | No of family members |  |  |  |
|  | Less than or equal to 4 family members | 13(28.3) | 20(43.5) | 33 |
|  | More than 4 family members | 33(71.7) | 26(56.5) | 59 |
| 8 | Under 5 children |  |  |  |
|  | Less than or equal to two children | 36(78.3)1 | 43(93.5) | 79 |
|  | More than 2 children | 10(21.7) | 3(6.5) | 30 |
| 9 | Years of experience |  |  |  |
|  | <= 5 years of experience | 23(50) | 15(32.6) | 38 |
|  | > 5 years of experience | 23(50) | 31(67.4) | 54 |
| 10 | Training |  |  |  |
|  | Less than or equal to 5 | 44(95.7) | 40(87) | 84 |
|  | More than 5 | 2(4.3) | 6(13) | 8 |

### The five-step process is as follows

#### Step 1: Review of literature

From the review of literature nearly 27 individual items under 19 major domains in Motivation scale and 18 individual items in job satisfaction scale were initially enlisted as enclosed in Supplementary File 1. Those items in each scale were first translated by 3 translators separately and then discrepancies were resolved and synthesized to single motivation and satisfaction questionnaires. It was reviewed again by the investigators and was distributed to the experts for content validation and translational errors through emails and as soft copy to the experts for their ratings as shown in Supplementary File 2.

#### Step 2: Internal validity of the questionnaire

In our study, the content validation technique (21) was adopted to ensure that each of the items intended to measure and is representative of the main objective i.e., motivation and job satisfaction. Here content validation was performed by an expert of 5 academic community medicine professionals and two Senior residents of Community and Family Medicine. Each expert was asked to rate each item in a scale of 1-Not relevant to 4-Strongly relevant. Those items which received score of 3 or 4 was indicated as relevant. Items with content validity I-CVI of 0.78 or higher for 6-10 experts were considered. In our study the Mean I-CVI for the motivation scale was found to be 0.63 and for the job satisfaction was found to be 0.82 as shown in Supplementary File 2.

#### Step 3: Pilot testing

A pilot study was conducted following alteration of the questionnaire and was administered to the CHWs excluding our study area of interest. Based on the feedback and the opinion of the participants and the investigators the questions were further modified, added, or removed and the repeat I-CVI was found to be 0.66 for motivation scale and 0.83 for the job satisfaction scale. The motivation questionnaire was shortlisted to 18 from 27 questions and Job satisfaction questionnaire was shortlisted to 15 from 18. The final modified questionnaire has been attached in Supplementary File 3.

#### Step 4: Exploratory factor analysis

This method was adopted to group the inter correlated variables under a separate component or a construct. The sample size was estimated using a rule of thumb rule where there should be at least 5 participants for every item. According to this rule for our 18 items of motivation questionnaire there must be 90 sample size and for 15 items of job satisfaction questionnaire there must be 75 sample size which were met. The questionnaire was administered to those who satisfied the inclusion criteria.

#### Step 5: Principal Component Analysis

Before the factor analysis, correlation matrix was constructed between the items of both the questionnaire in order to check the suitability of the data for factor analysis with the accepted value ranging from 0.30 to 0.85.

KMO (Kaiser Meyer Olkin test), test of sampling adequacy had a value 0.78 for motivation scale and 0.80 for job satisfaction indicating that the sample is adequate, however Bartlett’s test of sphericity had a p value of<0.001 for motivation and job satisfaction indicating further analysis.

##### For the motivation questionnaire

Using extraction method of PCA only the five components among the 18 have eigenvalues over 1.00, and together these explained over 61.92% of the total variability in the data. This led us to the conclusion that a 5 factor/component solution will probably be adequate as shown in Figure 1.

**Figure 1:**
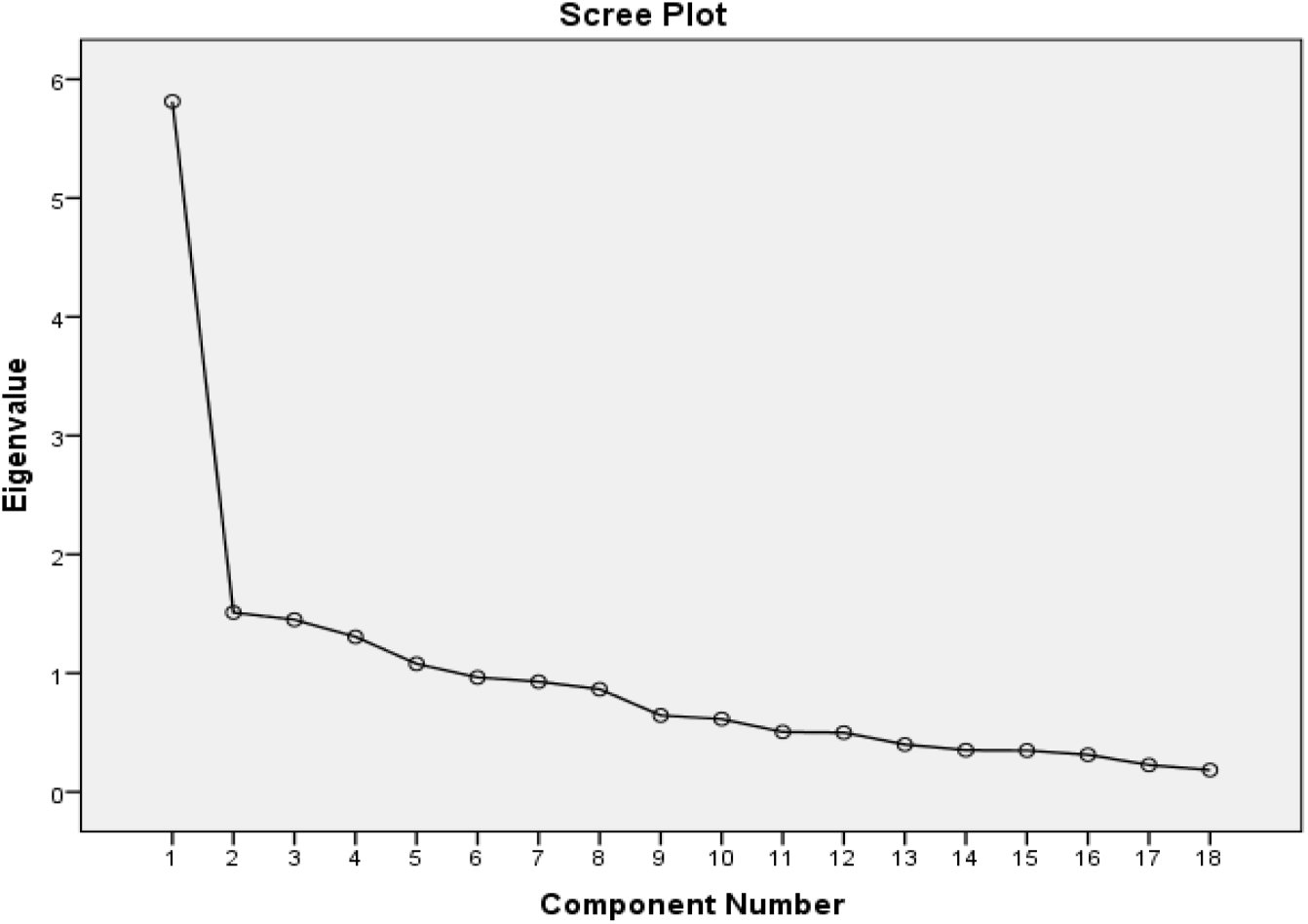
Screen plot displaying the components of Performance motivation scale.

##### For Job satisfaction questionnaire

Using extraction method of PCA only the three components among the 15 have eigenvalues over 1.00, and together these explained over 58.65% of the total variability in the data. This led us to the conclusion that a 3 factor/component solution will probably be adequate as shown by Scree plot in Figure 2.

**Figure 2:**
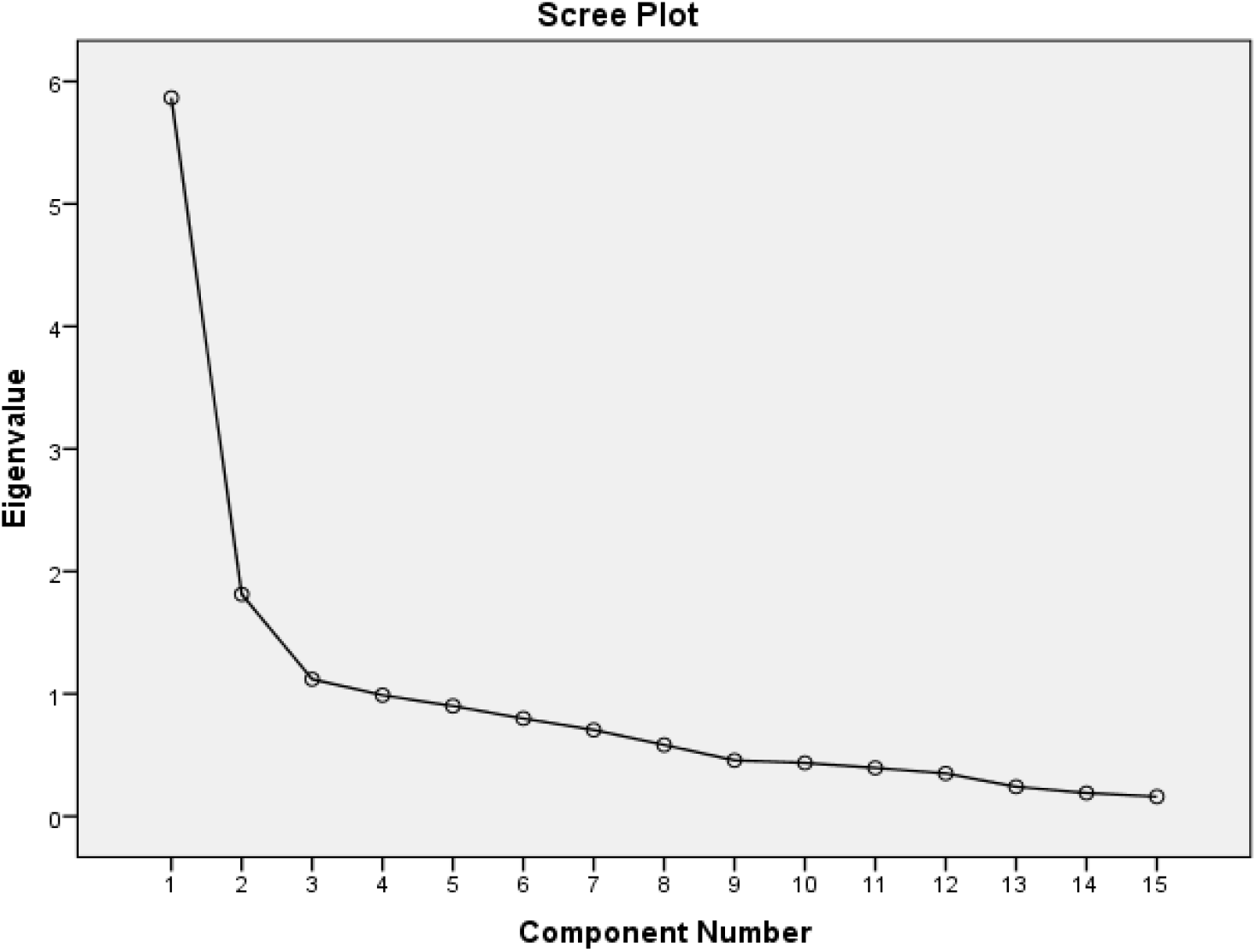
Screen plot displaying the components of Job satisfaction scale.

The factor loadings of all the 18 variables of motivation scale and 15 variables of job satisfaction questionnaire resulting from Varimax rotation are given in Supplementary File 4. The effect of rotation is to spread the importance equally between the rotated factors. For those variables which had occupied more than 1 component, they were allotted the respective components with value >0.5.

#### Step 5: Reliability Analysis

Reliability Analysis is defined as degree of consistency between different items in a construct. We had used Cronbach’s alpha to measure consistency where a value >=0.60 is considered reliable and acceptable (11).

Hence in the motivation questionnaire, the components 1 was accepted as their Cronbach’s was >0.8, in component 2 on deleting the variable PM3 alpha value was 0.624 and it was also accepted but the components 3, 4 and 5 were deleted as their Cronbach’s alpha was found to be and cannot be further manipulated the details of which are given in Supplementary File 4. While the components 1,2 and 3 of the job questionnaire was accepted as their Cronbach’ was >0.8.

### Summary of the assessment of performance motivation of CHWs as per the shortlisted questionnaire

The Finalized components of performance motivation included 11 questions under two components which were renamed as:

1. Intrinsic motivation component that included three questions,
2. Mixed component that included a combination of Individual, health system and community driven factors of motivation i.e., 8 questions and are displayed as below in Table 2.

**Table 2:** Finalized components of performance motivation scale with distribution of CHWs motivation score stratified by performance.

| Sl No | Variables | RLP Median (IQR) | RHP Median (IQR) | Total | Mann Whitney test |
| --- | --- | --- | --- | --- | --- |
| **Intrinsic Motivation component:** |  |  |  |  |  |
| 1 | I am confident enough to execute my responsibilities by myself | 4(4-5) | 4(4-5) | 4(4-5) | 0.130 |
| 2 | I am able to complete my daily tasks as I have scheduled, and I am able to spend time with my family. | 4(2-5) | 4(2-4) | 4(2-5) | 0.434 |
| 3 | I am able to handle difficult situations, solve problems by myself, feel emotionally and physically perfect on work. | 5(4-5) | 4(4-5) | 4(4-5) | 0.037 |
| **Mixed : (Individual +Health system Community driven factors of motivation)** |  |  |  |  |  |
| 4 | I am able to receive good number of trainings from my supervisor and able to obtain good knowledge | 5(4-5) | 4(4-5) | 5(4-5) | 0.870 |
| 5 | I am proud to be working for this health facility. | 4(4-5) | 4(4-5) | 4(4-5) | 0.308 |
| 6 | I am able to receive community's interest, acceptance and participation in the activities I perform | 5(4-5) | 4(4-5) | 4(4-5) | 0.459 |
| 7 | I want to improve community health | 4(4-5) | 4(3.75-5) | 4(4-5) | 0.293 |
| 8 | I am still interested in performing the social work even when existing social norms adversely impact the community health. | 4(3.5-5) | 4(4-5) | 4(4-5) | 0.200 |
| 9 | I am working with the idea that job is important and not merely for money alone. | 4(2.75-5) | 4(4-5) | 4(4-5) | 0.816 |
| 10 | I feel motivated to work hard | 5(4-5) | 5(4-5) | 4(4-5) | 0.035 |
| 11 | I have been given the freedom to move around freely in the community and express opinions and execute the responsibilities | 5(4-5) | 5(4-5) | 4(4-5) | 0.496 |
| **Overall score** |  | 4(4-5) | 4(4-5) | 4(4-5) |  |

Assessment of level of motivation using Likert scale showed that CHWs were well motivated with individual average score for each of the 11 questions above or equal to 4. Overall score was 4(4–5) among RLP CHWs and 4(4–5) among RHP CHWs.The component 1 in performance motivation scale was a complex interaction of the individual determinants like *(feeling of social responsibility and self-motivation*), Health system factors like *(involvement in trainings and Commitment in providing primary health care facilities)*, Community level factors like *(community participation and autonomy)*. Whereas the component 2 was predominantly based on health system factors like the workload and nature of job responsibility. Among the 11 questions, 2 items like “*I am able to handle difficult situations, solve problems by myself, feel emotionally and physically perfect on work”* (P value 0.037) and “*I feel motivated to work hard”* (P value 0.035) *were* found to be significantly associated on Mann Whitney test the details of which are given in Table 2.

### Summary of the assessment of job satisfaction of CHWs as per the shortlisted questionnaire

The Finalized components of job satisfaction included 15 questions under three components which were renamed as:

1. Health System component that included 5 questions,
2. Supervision and peer support component that included 7 questions,
3. Intrinsic job satisfaction component that included 3 questions and are displayed in Table 3.

**Table 3:** Finalized components of job satisfaction scale with distribution of CHWs job satisfaction score stratified by performance.

| Sl No | Variables | RLP Median (IQR) | RHP Median (IQR) | Total | P value Mann Whitney |
| --- | --- | --- | --- | --- | --- |
| **Health System component** |  |  |  |  |  |
| 1 | The set rules and regulations make it easy for me to do good job | 4(4-5) | 4(4-5) | 4(4-5) | 0.925 |
| 2 | I have adequate opportunities to develop my professional skills | 4(3.75-5) | 4(4-5) | 4(4-5) | 0.956 |
| 3 | I am provided with all trainings necessary for me to perform my job | 4(4-5) | 4(4-5) | 4(4-5) | 0.975 |
| 4 | I feel there is a sufficient workspace to do my job | 4(4-5) | 4(4-5) | 4(4-5) | 0.445 |
| 5 | My work is evaluated based on a fair system of performance standards. | 4.5(4-5) | 4(4-5) | 4(4-5) | 0.210 |
| **Supervision and the peer group component** |  |  |  |  |  |
| 6 | My work assignments are always clearly explained to me | 4(4-5) | 4(4-5) | 4(4-5) | 0.804 |
| 7 | I receive the right amount of support and guidance from my direct supervisor | 4(4-5) | 4(4-5) | 4(4-5) | 0.126 |
| 8 | I am appropriately recognized when I perform well at my regular work duties | 4(4-5) | 4(4-5) | 4(4-5) | 0.400 |
| 9 | I feel encouraged by my supervisor to offer suggestions and improvements | 4(4-5) | 4(4-5) | 4(4-5) | 0.852 |
| 10 | My department provides all the equipment, supplies and resources necessary for me to perform my duties | 4(4-5) | 4(4-5) | 4(4-5) | 0.391 |
| 11 | I have learnt many new job skills in this position | 4(3-4) | 4(4-5) | 4(4-5) | 0.004 |
| 12 | I have too much paperwork (reverse coded) | 2(1-3) | 2(1-2) | 1(1-2) | 0.303 |
| **Intrinsic Job satisfaction** |  |  |  |  |  |
| 13 | I feel I can easily communicate with members from all levels of this organization | 4(4-4) | 4(4-5) | 4(4-5) | 0.014 |
| 14 | I like doing the things I do at work | 4(4-5) | 4(4-5) | 4(4-5) | 0.074 |
| 15 | My work gives me a feeling of personal accomplishment | 4(4-5) | 4(4-5) | 4(4-5) | 0.066 |
| **Overall score** |  | 4(4-5) | 4(4-5) | 4(4-5) |  |

Assessment of level of job satisfaction using Likert scale from 1-5 displayed in Table 3 showed that CHWs were satisfied with their performance with individual average score for each of the 15 questions above or equal to 4. Overall median score was 4 (4–5) among RLP CHWs and 4(4-5) among RHP CHWs. The Component 1 in the job satisfaction scale explains those which are provided broadly by the Government under CHW program which leads to accomplishing job satisfaction. In component 2 job satisfaction tends to be contributed by the peer groups, supervisors, and the other workforce CHWs tend to work with while the final component 3 were those intrinsic factors of CHWs which provide job satisfaction. Among which items like “*I have learnt many new job skills in this position*” (P value 0.004) and “I *feel I can easily communicate with members from all levels”* (P value 0.014) were found to be significantly associated on Mann Whitney test. It is to be noted that one variable among 15 variables i.e. “I have too much paperwork” has been reverse coded (1-strong agreement to 5-strong disagreement) to minimize ascertainment bias.

## Discussion

Certain cross-cultural adaptations were made in our literature review as per the commonly used process of Beaton et al.(22) comprising of idiomatic and experiential equivalence (23). Also, the description of domains and the identification of items under each domain was based on deductive method or “logical partitioning” as explained by Boateng et al (24) where existing scales were assessed and grouped into final questionnaire unlike the inductive method that involves generation of items from the individuals’ responses. The I-CVI was 0.66 for motivation and 0.83 for job satisfaction that could be considered for 6-10 experts (11). Some authors use KMO sampling adequacy test to ensure adequate sample size (23).In our study, the sampling adequacy as accomplished by KMO values were 0.78 and 0.80 for motivation and job satisfaction. However, researchers use minimum 2 to maximum 20 people per item to estimate the sample size as arbitrary (23). It is said that larger the sample size, more stable would be the factor loadings and replicable factors to generalize the results (23).

The Explanatory factor analysis had extracted 11 questions in motivation scale and 15 questions in job satisfaction scale. The constructs showed good internal consistency with Cronbach’s alpha ranging from 0.62-0.88. Similar study (6) done in Orissa, enlisted 16 parameters of performance motivation but the information on the validity, reliability and factor analysis were not mentioned. Another study (5) in Haryana included a 23-item questionnaire which was pretested in the field among CHWs but lacked information on the validity and consistency of the questionnaire with unclear information on the conclusion of the constructs. Similar study (25) on job satisfaction administered in Iran contained 8 aspects of questions with inadequate validation, good consistency of 0.87 with no mention on factor analysis. Findings from other study (13) conducted in low-income settings like India using a 20-component questionnaire included information on convergent validity with good consistency of >0.70 and extraction of 3 factors by factor analysis that focussed on relation with the supervisors, co-workers, and job satisfaction.

The component 1 in performance motivation scale of our study was a complex interaction of the individual determinants, health system and community level factors whereas the component 2 was predominantly based on health system factors. Hence proves the fact that motivation does not solely depend on certain specific factor rather results from interactions with various other factors which was contradictory to the findings of Gopalan et.al (6) that found that CHWs were better motivated with individual and community factors. These findings in our study might have resulted as CHWs were intrinsically motivated and had received adequate support from the peers and the community. The current study observed that CHWs had significant intrinsic feeling of motivation to perform better and perceived that they were self-efficacious which was similar to this study (26). However, the components stratification has not been observed in other studies as compared to ours.

Similarly, the Component 1 in the job satisfaction scale explains those which are provided broadly by the Government under CHW programme which leads to accomplishing job satisfaction. In component 2 job satisfaction tends to be contributed by the peer groups, supervisors and the other workforce CHWs tend to work with. The final component 3 were those intrinsic factors of CHWs which provides job satisfaction. Thus, job satisfaction of CHWs tend to depend primarily on the intrinsic factor which was in turn influenced by the workplace factors and the amendments implemented at national level for the programme. Unlike other studies (5,6) that focused on the overall mean score of motivation and job satisfaction and the association with their determinants, our study was novel in associating every finalized item of both the scales with their performance-based incentives (RHP/RLP).

## Strengths

This was a novel study focusing on the least explored yet critical determinants of CHWs performance. Unlike other studies in India our study covered a wholesome picture from content validation up to reliability analysis. Self-administered questionnaire using mobile based app was employed to prevent interviewer bias, reduce the errors during data entry and facilitate the monitoring of data.

## Limitations

However, due to the purposive sampling technique employed, the generalizability of results cannot be done. Further since we did not have enough sample size i.e., independent cluster of participants to partition the data for confirmatory analysis, we were not able to validate the constructs generated. Also, our tools lacked number of reverse coded questions and therefore the possibility of the ascertainment bias cannot be ruled out in the motivation and the job satisfaction questionnaire.

## Conclusion

The motivation and job satisfaction scale has I-CVI ranging from 0.6-0.8 and good internal consistency with Cronbach’s alpha > 0.8 including constructs from various components. The overall median (IQR) score of both RLP and RHP CHWs being 4(4–5) in both the questionnaires suggesting that they are intrinsically motivated and were satisfied with their performance as CHWs. Further research would be planned to validate the constructs using confirmatory factor analysis.

## Data Availability

All data produced in the present study are available upon reasonable request to the authors

## Abbreviations

ASHA: Accredited Social Health Activists
CHC: Community Health Centre
CHWs: Community Health workers
I-CVI: Item Content validity index
IQR: Interquartile range
NHM: National Health Mission
ODK: Open Data Kit
ONA: Organizational network analysis
PHC: Primary Health Centre
RHP: Relatively High Performing
RLP: Relatively Low Performing
XLS: eXcel Spreadsheet

## Acknowledgement

Our sincere thanks to Dr. Deepti Dabar for her valuable guidance and feedback during the process of protocol writing and submission.

We thank our Head of the department, Prof (Dr.) Arun Kokane for his support that had helped us to mobilize the CHWs during our study at various settings.

We thank all the Senior residents, Junior Residents and the interns who helped us during the data collection period and for their cooperation during the study.

## Supplementary Files

**Supplementary File 1:** supplementary table 1 and 2.

**Supplementary File 2:** supplementary table 3 and 4.

**Supplementary File 3:** supplementary table 5 to 8.

**Supplementary File 4:** supplementary table 9 to 14.

