## Supplementary File 1 for "Development of tool for assessment of performance motivation and job satisfaction among the Community Health Workers of Central India"

### Supplementary table 1: Level of performance motivation among the CHWs (27 scale questionnaire).

|  | Variables | | |  | | Strongly Disagree | | | Disagree | | Neutral | | Agree | | Strongly agree |
| --- | --- | --- | --- | --- | --- | --- | --- | --- | --- | --- | --- | --- | --- | --- | --- |
|  | Health system level: | | | | | | | | | | | | | | |
|  | Nature of responsibilities: | | | I am confident enough to execute my responsibilities by myself | | 1 | | | 2 | | 3 | | 4 | | 5 |
|  | Workload: | | | I am able to complete my daily tasks as I have scheduled, and I am able to spend time with my family.  I feel emotionally drained at the end of the day | | 1  1 | | | 2  2 | | 3  3 | | 4  4 | | 5  5 |
|  | Incentive: | | | I feel adequate with the incentives I receive.  I do this job as I am getting paid for it | | 1  1 | | | 2  2 | | 3  3 | | 4  4 | | 5  5 |
|  | Health care infrastructure | | | I am satisfied on the quality of existing infrastructure, communication options and commodities. | | 1 | | | 2 | | 3 | | 4 | | 5 |
|  | Work modality | | | I am satisfied with the existing recording and reporting system  I always complete my tasks efficient and correctly | | 1  1 | | | 2  2 | | 3  3 | | 4  4 | | 5  5 |
|  | Training: | | | I am able to receive good number of trainings from my supervisor and able to obtain good knowledge. | | 1 | | | 2 | | 3 | | 4 | | 5 |
| Peer support | | | I am having good moral support from my peer group. | | 1 | | | 2 | | 3 | | 4 | | 5 | |
|  | | | I can rely on my colleagues at work | | 1 | | | 2 | | 3 | | 4 | | 5 | |
| Organization Commitment | | | I am proud to be working for those health facility | | 1 | | | 2 | | 3 | | 4 | | 5 | |
|  | | | I feel very committed to this health facility | | 1 | | | 2 | | 3 | | 4 | | 5 | |
| Community level: | | | | | | | | | | | | | | | |
| Community participation | | | I am able to receive community ‘s interest, acceptance and participation in the activities I perform. | | 1 | | | 2 | | 3 | | 4 | | 5 | |
| Improve  community health | | | I want to improve my community health | | 1 | | | 2 | | 3 | | 4 | | 5 | |
| Individual level: | | | | | | | | | | | | | | | |
| Social responsibility and altruism: | | | I am still interested in performing the social work even when existing social norms adversely impact the community health. | | 1 | | | 2 | | 3 | | 4 | | 5 | |
| Self-efficacy: | | | I am able to handle difficult situations, solve problems by myself, feel emotionally and physically perfect on work. | | 1 | | | 2 | | 3 | | 4 | | 5 | |
| Self-motivation: | | | I am working with the idea that job is important and not merely for money alone. | | 1 | | | 2 | | 3 | | 4 | | 5 | |
| General motivation | | | I feel motivated to work hard | | 1 | | | 2 | | 3 | | 4 | | 5 | |
|  | Job security | I am doing this job as it provides long term security for me | | | | | 1 | 2 | | 3 | | 4 | | 5 | |
|  | Self-development | I wanted to eventually work elsewhere and knew that field experience was required | | | | | 1 | 2 | | 3 | | 4 | | 5 | |
|  |  | I hoped to gain skills that would enable me to work as a health worker | | | | | 1 | 2 | | 3 | | 4 | | 5 | |
|  |  | I expected the experience will enhance my communication skills | | | | | 1 | 2 | | 3 | | 4 | | 5 | |
|  | Value and recognition | My family and friends encouraged me apply for this job | | | | | 1 | 2 | | 3 | | 4 | | 5 | |
|  |  | I knew of the other ASHAs who were respected in the community | | | | | 1 | 2 | | 3 | | 4 | | 5 | |
|  | Individual + community+ Health system level: | | | | | | | | | | | | | | |
|  | Recognition | My performance is being valued and appreciated by my family, community and system | | | | | 1 | 2 | | 3 | | 4 | | 5 | |
|  | Autonomy | I have been given the freedom to move around freely in the community and express opinions and execute the responsibilities. | | | | | 1 | 2 | | 3 | | 4 | | 5 | |

**Supplementary table 2: Level of job satisfaction among the CHWs (18 scale questionnaire).**

| *Questions* | *Strongly disagree* | *Disagree* | *Neutral* | *Agree* | *Strongly Agree* |
| --- | --- | --- | --- | --- | --- |
| I feel encouraged by my supervisor to offer suggestions and improvements | 1 | 2 | 3 | 4 | 5 |
| My Co-workers and I work well together | 1 | 2 | 3 | 4 | 5 |
| The set rules and regulations make it easy for me to do good job | 1 | 2 | 3 | 4 | 5 |
| I have adequate opportunities to develop my professional skills | 1 | 2 | 3 | 4 | 5 |
| I have an accurate written job prescription | 1 | 2 | 3 | 4 | 5 |
| My work assignments are always clearly explained to me | 1 | 2 | 3 | 4 | 5 |
| I receive the right amount of support and guidance from my direct supervisor | 1 | 2 | 3 | 4 | 5 |
| I am provided with all trainings necessary for me to perform my job | 1 | 2 | 3 | 4 | 5 |
| I am appropriately recognized when I perform well at my regular work duties | 1 | 2 | 3 | 4 | 5 |
| I have too much paperwork | 1 | 2 | 3 | 4 | 5 |
| I am satisfied with my chances for promotion | 1 | 2 | 3 | 4 | 5 |
| My work is evaluated based on a fair system of performance standards. | 1 | 2 | 3 | 4 | 5 |
| My department provides all the equipment, supplies and resources necessary for me to perform my duties | 1 | 2 | 3 | 4 | 5 |
| I have learnt many new job skills in this position | 1 | 2 | 3 | 4 | 5 |
| I feel I can easily communicate with members from all levels of this organization | 1 | 2 | 3 | 4 | 5 |
| I like doing things I do at work | 1 | 2 | 3 | 4 | 5 |
| My work gives me a feeling of personal accomplishment | 1 | 2 | 3 | 4 | 5 |
| I feel there is a sufficient workspace to do my job | 1 | 2 | 3 | 4 | 5 |
