## Supplementary File 2 for "Development of tool for assessment of performance motivation and job satisfaction among the Community Health Workers of Central India"

**Supplementary table 3: Content validation of performance motivation questionnaire**

Total no of experts: 7

(Neutral/strongly agree/agree=1 & Strongly disagree/disagree= 0)

|  | A | B | C | D | E | F | G | No  experts in agreement | I-CVI |
| --- | --- | --- | --- | --- | --- | --- | --- | --- | --- |
| PM1 | 1 | 0 | 0 | 1 | 0 | 0 | 1 | 3 | 0.4 |
| PM2 | 1 | 0 | 0 | 1 | 0 | 0 | 0 | 2 | 0.3 |
| PM3 | 1 | 0 | 1 | 1 | 1 | 0 | 1 | 5 | 0.7 |
| PM4 | 1 | 1 | 1 | 1 | 1 | 0 | 1 | 6 | 0.9 |
| PM5 | 0 | 0 | 0 | 0 | 0 | 1 | 1 | 2 | 0.3 |
| PM6 | 1 | 0 | 0 | 1 | 1 | 0 | 0 | 3 | 0.4 |
| PM7 | 1 | 0 | 0 | 1 | 1 | 1 | 1 | 4 | 0.6 |
| PM8 | 1 | 0 | 1 | 0 | 1 | 1 | 1 | 5 | 0.7 |
| PM9 | 1 | 1 | 0 | 0 | 1 | 1 | 1 | 5 | 0.7 |
| PM10 | 1 | 1 | 0 | 0 | 0 | 0 | 1 | 3 | 0.4 |
| PM11 | 1 | 1 | 1 | 0 | 0 | 1 | 1 | 5 | 0.7 |
| PM12 | 0 | 1 | 1 | 1 | 0 | 1 | 1 | 5 | 0.7 |
| PM13 | 1 | 1 | 1 | 0 | 0 | 1 | 1 | 5 | 0.7 |
| PM14 | 0 | 1 | 0 | 1 | 0 | 0 | 1 | 3 | 0.4 |
| PM15 | 0 | 1 | 1 | 1 | 0 | 1 | 1 | 5 | 0.7 |
| PM16 | 1 | 1 | 1 | 0 | 1 | 1 | 1 | 6 | 0.9 |
| PM17 | 1 | 0 | 1 | 1 | 1 | 1 | 1 | 6 | 0.9 |
| PM18 | 1 | 0 | 1 | 1 | 1 | 1 | 1 | 6 | 0.9 |
| PM19 | 1 | 0 | 1 | 0 | 0 | 1 | 1 | 4 | 0.6 |
| PM20 | 1 | 1 | 0 | 1 | 0 | 1 | 1 | 5 | 0.7 |
| PM21 | 0 | 0 | 1 | 0 | 0 | 1 | 0 | 2 | 0.3 |
| PM22 | 0 | 1 | 0 | 1 | 1 | 0 | 0 | 3 | 0.4 |
| PM23 | 1 | 1 | 0 | 0 | 1 | 1 | 1 | 5 | 0.7 |
| PM24 | 1 | 0 | 0 | 1 | 0 | 1 | 0 | 3 | 0.4 |
| PM25 | 0 | 1 | 0 | 1 | 0 | 1 | 1 | 4 | 0.6 |
| PM26 | 0 | 1 | 1 | 1 | 1 | 1 | 1 | 6 | 0.9 |
| PM27 | 0 | 1 | 0 | 1 | 1 | 1 | 1 | 5 | 0.7 |
| Total | 0.67 | 0.55 | 0.48 | 0.63 | 0.51 | 0.88 | 0.81 |  | 16.9 |

Mean I-CVI= 0.63

Mean expert proportion: 0.63

**Supplementary table 4: Content validation of job satisfaction questionnaire**

|  | A | B | C | D | E | F | G | No experts  in agreement | I-CVI |
| --- | --- | --- | --- | --- | --- | --- | --- | --- | --- |
| S1 | 1 | 1 | 1 | 1 | 1 | 1 | 1 | 7 | 1 |
| S2 | 1 | 1 | 0 | 1 | 0 | 1 | 1 | 5 | 0.7 |
| S3 | 1 | 1 | 1 | 1 | 0 | 1 | 0 | 5 | 0.7 |
| S4 | 1 | 1 | 1 | 1 | 1 | 0 | 0 | 5 | 0.7 |
| S5 | 0 | 1 | 0 | 0 | 1 | 0 | 0 | 2 | 0.3 |
| S6 | 0 | 1 | 1 | 1 | 1 | 1 | 1 | 6 | 0.9 |
| S7 | 1 | 1 | 1 | 1 | 1 | 1 | 1 | 7 | 1 |
| S8 | 1 | 1 | 1 | 1 | 1 | 1 | 1 | 7 | 1 |
| S9 | 1 | 1 | 1 | 1 | 1 | 1 | 1 | 7 | 1 |
| S10 | 1 | 0 | 1 | 1 | 1 | 1 | 1 | 6 | 0.9 |
| S11 | 1 | 1 | 1 | 1 | 1 | 1 | 1 | 7 | 1 |
| S12 | 1 | 1 | 1 | 1 | 1 | 1 | 1 | 7 | 1 |
| S13 | 0 | 1 | 1 | 1 | 1 | 1 | 1 | 6 | 0.9 |
| S14 | 0 | 1 | 0 | 1 | 1 | 1 | 1 | 5 | 0.7 |
| S15 | 0 | 1 | 1 | 1 | 1 | 1 | 1 | 6 | 0.9 |
| S16 | 0 | 1 | 1 | 0 | 0 | 1 | 1 | 4 | 0.6 |
| S17 | 1 | 1 | 1 | 0 | 0 | 1 | 1 | 5 | 0.7 |
| S18 | 1 | 1 | 0 | 1 | 1 | 1 | 1 | 6 | 0.9 |
| Total | 0.67 | 0.94 | 0.78 | 0.83 | 0.78 | 0.83 | 0.88 |  | 14.9 |

Mean I-CVI= 0.82

Mean expert proportion: 0.82
