## Supplementary File 3 for "Development of tool for assessment of performance motivation and job satisfaction among the Community Health Workers of Central India"

**Supplementary table 6: Finalized questionnaire of Job satisfaction: (15 questions)**

| *Questions* | *Strongly disagree* | *Disagree* | *Neutral* | *Strongly Agree* | *Agree* |
| --- | --- | --- | --- | --- | --- |
| I feel encouraged by my supervisor to offer suggestions and improvements  My Co-workers and I work well together  The set rules and regulations make it easy for me to do good job  I have adequate opportunities to develop my professional skills  I have an accurate written job prescription  My work assignments are always clearly explained to me  I receive the right amount of support and guidance from my direct supervisor  I am provided with all trainings necessary for me to perform my job | 1  1  1  1  1  1  1  1 | 2  2  2  2  2  2  2  2 | 3  3  3  3  3  3  3  3 | 4  4  4  4  4  4  4  4 | 5  5  5  5  5  5  5  5 |
| I am appropriately recognized when I perform well at my regular work duties.  I have too much paperwork | 1  1 | 2  2 | 3  3 | 4  4 | 5  5 |
| I am satisfied with my chances for promotion | 1 | 2 | 3 | 4 | 5 |
| My work is evaluated based on a fair system of performance standards. | 1 | 2 | 3 | 4 | 5 |
| My department provides all the equipment, supplies and resources necessary for me to perform my duties | 1 | 2 | 3 | 4 | 5 |
| I have learnt many new job skills in this position | 1 | 2 | 3 | 4 | 5 |
| I feel I can easily communicate with  members from all levels of this organization | 1 | 2 | 3 | 4 | 5 |

**Supplementary table 7 : Distribution of responses of Motivation score stratified by the ASHAs performance (N=92)**

| **Variables** | **Performance**  **RHP (N=46)**  **RLP (N=46)** | **Strongly disagree** | **Disagree** | **Neutral** | **Agree** | **Strongly agree** |
| --- | --- | --- | --- | --- | --- | --- |
| **PM1** | **RHP** | 0 | 2 | 2 | 21 | 21 |
|  | **RLP** | 5 | 2 | 2 | 21 | 16 |
| **PM2** | **RHP** | 3 | 9 | 6 | 18 | 9 |
|  | **RLP** | 4 | 9 | 2 | 17 | 14 |
| **PM3** | **RHP** | 5 | 8 | 3 | 19 | 10 |
|  | **RLP** | 5 | 9 | 2 | 18 | 12 |
| **PM4** | **RHP** | 9 | 15 | 5 | 9 | 8 |
|  | **RLP** | 9 | 12 | 5 | 11 | 9 |
| **PM5** | **RHP** | 3 | 4 | 11 | 18 | 9 |
|  | **RLP** | 7 | 12 | 4 | 8 | 15 |
| **PM6** | **RHP** | 5 | 2 | 0 | 27 | 12 |
|  | **RLP** | 4 | 3 | 0 | 16 | 23 |
| **PM7** | **RHP** | 1 | 2 | 0 | 22 | 21 |
|  | **RLP** | 6 | 1 | 5 | 16 | 18 |
| **PM8** | **RHP** | 1 | 0 | 2 | 28 | 13 |
|  | **RLP** | 4 | 3 | 1 | 9 | 29 |
| **PM9** | **RHP** | 1 | 1 | 1 | 22 | 21 |
|  | **RLP** | 3 | 5 | 2 | 10 | 26 |
| **PM10** | **RHP** | 2 | 2 | 1 | 26 | 14 |
|  | **RLP** | 3 | 7 | 1 | 15 | 20 |
| **PM11** | **RHP** | 1 | 0 | 2 | 21 | 22 |
|  | **RLP** | 5 | 2 | 0 | 20 | 19 |
| **PM12** | **RHP** | 0 | 1 | 3 | 22 | 20 |
|  | **RLP** | 2 | 2 | 1 | 16 | 25 |
| **PM13** | **RHP** | 1 | 6 | 4 | 23 | 12 |
|  | **RLP** | 4 | 2 | 2 | 20 | 17 |
| **PM14** | **RHP** | 0 | 3 | 3 | 18 | 22 |
|  | **RLP** | 3 | 7 | 1 | 16 | 18 |
| **PM15** | **RHP** | 3 | 4 | 2 | 21 | 14 |
|  | **RLP** | 6 | 5 | 1 | 18 | 16 |
| **PM16** | **RHP** | 0 | 0 | 1 | 20 | 25 |
|  | **RLP** | 3 | 4 | 2 | 19 | 18 |
| **PM17** | **RHP** | 1 | 4 | 1 | 19 | 21 |
|  | **RLP** | 2 | 4 | 3 | 16 | 21 |
| **PM18** | **RHP** | 1 | 1 | 4 | 16 | 24 |
|  | **RLP** | 3 | 2 | 2 | 18 | 21 |

**Supplementary table 8 : Distribution of responses of Job satisfaction score stratified by the ASHAs performance (N=92)**

| **Variables** | **Performance**  **RHP (N=46)**  **RLP (N=46)** | **Strongly disagree** | **Disagree** | **Neutral** | **Agree** | **Strongly agree** |
| --- | --- | --- | --- | --- | --- | --- |
| **S1** | **RHP** | 2 | 0 | 4 | 23 | 17 |
|  | **RLP** | 3 | 3 | 0 | 22 | 18 |
| **S2** | **RHP** | 1 | 3 | 6 | 26 | 10 |
|  | **RLP** | 3 | 6 | 2 | 23 | 12 |
| **S3** | **RHP** | 2 | 2 | 1 | 27 | 14 |
|  | **RLP** | 1 | 2 | 2 | 27 | 14 |
| **S4** | **RHP** | 3 | 0 | 0 | 23 | 20 |
|  | **RLP** | 3 | 2 | 0 | 24 | 17 |
| **S5** | **RHP** | 0 | 1 | 1 | 29 | 15 |
|  | **RLP** | 1 | 3 | 0 | 19 | 23 |
| **S6** | **RHP** | 1 | 2 | 2 | 27 | 12 |
|  | **RLP** | 4 | 2 | 1 | 26 | 13 |
| **S7** | **RHP** | 2 | 0 | 3 | 23 | 18 |
|  | **RLP** | 4 | 1 | 4 | 24 | 12 |
| **S8** | **RHP** | 1 | 4 | 2 | 24 | 14 |
|  | **RLP** | 5 | 2 | 3 | 24 | 12 |
| **S9** | **RHP** | 2 | 4 | 3 | 23 | 14 |
|  | **RLP** | 2 | 3 | 4 | 22 | 15 |
| **S10** | **RHP** | 0 | 3 | 0 | 24 | 19 |
|  | **RLP** | 1 | 4 | 1 | 23 | 16 |
| **S11** | **RHP** | 0 | 1 | 1 | 23 | 18 |
|  | **RLP** | 1 | 7 | 4 | 24 | 10 |
| **S12** | **RHP** | 1 | 3 | 5 | 21 | 16 |
|  | **RLP** | 3 | 4 | 6 | 20 | 13 |
| **S13** | **RHP** | 0 | 1 | 3 | 20 | 21 |
|  | **RLP** | 2 | 2 | 4 | 26 | 10 |
| **S14** | **RHP** | 0 | 1 | 3 | 20 | 19 |
|  | **RLP** | 4 | 2 | 2 | 25 | 13 |
| **S15** | **RHP** | 2 | 1 | 2 | 22 | 19 |
|  | **RLP** | 5 | 3 | 2 | 24 | 12 |
