## Supplementary File 4 for "Development of tool for assessment of performance motivation and job satisfaction among the Community Health Workers of Central India"

### Supplementary table 9: Factor loadings of all the variables of Performance motivation scale

|  | Component 1 | Component 2 Component | 3 Component 4 Component 5 |
| --- | --- | --- | --- |
| PM9 | .746 | .309 |  |
| PM11 | .727 |  |  |
| PM15 | .684 | .302 |  |
| PM8 | .662 |  |  |
| PM16 | .650 | .332 |  |
| PM12 | .611 | .425 |  |
| PM18 | .590 | .366 |  |
| PM13 | .516 |  | .346 |
| PM2 |  | .731 |  |
| PM1 | .441 | .624 |  |
| PM3 |  | .606 |  |
| PM14 | .331 | .450 |  |
| PM10 |  | .842 |  |
| PM17 | .381 | .712 |  |
| PM5 |  |  | .824 |
| PM4 | -.371 |  | .792 |
| PM6 |  |  | .881 |
| PM7 | .334 | .414 | .450 |

Note PM- Performance motivation

**Supplementary table 10: Grouping of the variables of performance motivation scale to the different components**

| Component | Variables included |
| --- | --- |
| 1 | PM9, 11, 15, 8,16, 12, 18 and 13. |
| 2 | PM2, 1, 3 and 14 |
| 3 | PM10, PM17 |
| 4 | PM4, PM5 |
| 5 | PM6, PM7 |

### Supplementary table 11: Factor loadings of all the variables of job satisfaction scale

|  |  | Component 1 | | Component 2 | | Component 3 |
| --- | --- | --- | --- | --- | --- | --- |
|  | S12 | .757 | |  | |  |
|  | S7 | .751 | | .387 | |  |
|  | S10 | .666 | | .315 | | .351 |
| S9 | | .592 | .496 | |  | |
| S4 | | .569 | .475 | |  | |
| S5 | | .554 |  | |  | |
| S1 | | .418 | .328 | |  | |
| S3 | |  | .809 | |  | |
| S8 | | .314 | .658 | | .328 | |
| S6 | | .450 | .612 | |  | |
| S2 | |  | .510 | |  | |
| S15 | |  | .488 | | .397 | |
| S14 | |  |  | | .816 | |
| S11 | |  |  | | .807 | |
| S13 | |  |  | | .787 | |

Note: S - Satisfaction

**Supplementary table 12: Grouping of the variables of the performance motivation scale to several components**

| Component | Variables included |
| --- | --- |
| 1 | S12, S7, S10, S9, S4, S5, S1 |
| 2  3 | S3, S8, S6, S2, S15  S14, S11, S13 |

**Supplementary table 13: Internal Consistency of various constructs of motivation questionnaire**

|  | Component | | Variables included | | Cronbach’s alpha |
| --- | --- | --- | --- | --- | --- |
|  | 1 | | PM9, 11, 15, 8,16, 12, 18 and 13. | | 0.843 |
|  | 2 | | PM2, 1, 3 and 14 | | 0.523 |
| 3 | | PM10, PM17 | | 0.564 | |
| 4 | | PM4, PM5 | | 0.510 | |
| 5 | | PM6, PM7 | | 0.515 | |
|  | | Overall, for 18 variables | | 0.822 | |

**Supplementary table 14: Internal Consistency of various constructs of job satisfaction questionnaire**

| Component | Variables included | Cronbach’s alpha |
| --- | --- | --- |
| 1 | S12, 7, 10,9,4,5 and 1 | 0.801 |
| 2 | S3, S8, S6, S2, S15 | 0.732 |
| 3 | S14, S11 and S13 | 0.788 |
|  | Overall, for 15 variables | 0.883 |
